# Age Matters: COVID-19 Prevalence in a Vaping Adolescent Population – An Observational Study

**DOI:** 10.1101/2020.07.03.20146035

**Authors:** Nitin Tandan, Manjari Rani Regmi, Ruby Maini, Abdisamad M. Ibrahim, Cameron Koester, Odalys Estefania Lara Garcia, Priyanka Parajuli, Mohammad Al-Akchar, Mukul Bhattarai, Abhishek Kulkarni, Robert Robinson

**Affiliations:** Department of Internal Medicine, Southern Illinois University School of Medicine, Springfield, IL, USA; Division of Cardiology, Southern Illinois University School of Medicine, Springfield, IL, USA

**Keywords:** COVID-19, Vaping, E-cigarettes, Adolescent, Prevalence

## Abstract

**Background:** Currently, there is limited or no data demonstrating that vaping is associated with increased transmission or prevalence of coronavirus disease-2019 (COVID-19). Our study aims to investigate the relationship of vaping with the prevalence of COVID-19 infection across the United States and in the District of Columbia.

**Methods:** COVID-19 case counts by state and the District of Columbia were obtained via the Worldometers website on 04/30/2020. Prevalence of COVID-19 cases per 100,000 residents were calculated using estimated 2019 population data from the US Census Department. Age ranges analyzed were: high school age children, Ages 18-24, Ages 25-44, and Ages 45-65. Spearman correlation analysis was conducted to determine if the rate of vaping was correlated with a higher prevalence of COVID-19 cases per 100,000 population.

**Findings:** The Spearman correlation analysis demonstrated that persons vaping between 18 years and 24 years of age had a correlation coefficient of 0.278 with prevalence of COVID-19 infection (p=0.048). Vaping high school students had a correlation coefficient of 0.153 with prevalence of COVID-19 (p=0.328). Persons vaping in the age group 25-45 years had a correlation coefficient of 0.101 in association to COVID-19 prevalence (p=0.478). And finally, persons vaping between the age 45-65 years old had a correlation coefficient 0.130 with respect to COVID-19 prevalence (p=0.364).

**Interpretation:** Increased COVID-19 prevalence is associated with vaping in the adolescent population between ages 18 and 24. Further prospective studies need to be performed in order investigate the severity of outcomes of vaping in association with COVID-19 infection.

**Funding:** Nothing to disclose.

## Introduction

Severe acute respiratory syndrome coronavirus-2 (SARS-CoV-2) is the novel coronavirus detected in Wuhan, China in December 2019, known to cause coronavirus disease 2019 (COVID-19). Since its first detection, it quickly escalated to become a pandemic affecting more than four million people across the world. To date, more than 1.85 million cases have been diagnosed with approximately 109,000 confirmed deaths in the United States (1). Worldwide, approximately 6.85 million cases have been confirmed with reported an approximate death total of 398,000 persons (2). Viruses from the Coronavirus family that have contributed to endemics include but are not limited to severe acute respiratory syndrome coronavirus (SARS-CoV) and Middle East respiratory syndrome coronavirus (MERS-CoV) (2). Coronaviruses are from the family of Coronaviridae. Initially thought to be transmitted through bats, it was later identified that person to person transmission is possible. Based upon the current understanding, coronavirus is transmitted mainly through respiratory droplets, aerosols, fomites, and body fluids (3). Aerosols (0.3-100 mm) containing virus can remain in the environment and even transmit the virus from 100 meters or more compared to droplets (1-5 mm) which transfer 1 to 2 meters from the source of infection (4). Therefore, hospitalized patients who are strongly suspected and/or positive for COVID-19 and have strong suspicions are isolated with airborne precautions in a negative air-pressure room to limit the transmission. However, we now know that a person can transmit the virus even before they present symptoms or that they can even be asymptomatic carriers. For those transmissions, physical distancing and avoiding of public places are highly encouraged among healthy individuals.

The prevalence of electronic cigarette use, or “vaping”, has been increasing in the United States especially in adolescents since first introduced in 2006. Vaping is the inhalation of an aerosol or vapor through a device such as electronic cigarettes. These products may or may not contain nicotine, marijuana, or other drugs (5). Some studies demonstrate that vaping can emit airborne particulate material that may be associated with passive smoking (6). It is unclear if vaping can lead to the emission of particles that can be inhaled by nearby persons. Currently, there is limited or no data thus far demonstrating that vaping is associated with increased transmission or prevalence of COVID-19. Our study aims to investigate the relationship of vaping with the prevalence of COVID-19 infection across the United States and in the District of Columbia.

## Methods

COVID-19 case counts by state and the District of Columbia were obtained via the Worldometers website on 04/30/2020 at 1300 GMT (7).

Prevalence of COVID-19 cases per 100,000 residents were calculated using estimated 2019 population data from the US Census Department (7).

E-cigarette use data by state and the District of Columbia was obtained via the CDC State Tobacco Activities Tracking and Evaluation (STATE) system.

Values by age range from the most recent survey of a state were used for analysis (8). Age ranges analyzed were: high school age children, Ages 18-24, Ages 25-44, and Ages 45-65. Vaping prevalence data for the high school age range was not available in the CDC STATE system for Connecticut, Florida, Georgia, Minnesota, New Jersey, Ohio, Oregon, and Washington.

Spearman correlation analysis was conducted to determine if the rate of vaping was correlated with a higher prevalence of COVID-19 cases per 100,000 population. Statistical analysis was conducted with SPSS version 26. P values are two sided. P values less than 0.05 were considered statistically significant.

No direct patient care or contact was undertaken while performing this observational study, thus this study was exempted from Institutional Review Board oversight.

### a.) Evidence before this study

Current evidence reported by the CDC demonstrated the significant devastation attributed to the COVID-19 infection. Over 4 million cases were diagnosed as of 05/13/2020 and over 700,000 deaths were reported in the span of 4 months since the origin of the infection, proposed to be at the end of December in 2019. The authors performed a thorough search and review of the available data in CDC, Scopus, PubMed, Journal of the American Medical Association, the Lancet, and New England Journal of Medicine. The literature assessed provided invaluable knowledge regarding the characteristics of COVID-19 as well as the toxic effects of vaping. With the help of these studies, we were able to perform an unique investigation correlating vaping and COVID-19.

### b.) Added value of this study

We present the first original investigation of associations of COVID-19 and vaporizer use (“vaping”). We used the public data available from Worldometers as well as from CDC regarding estimated population sized per state and regarding prevalence of COVID-19 and vaping across the 50 United States. We used a Spearman correlation analysis to assess correlation of vaping and COVID-19 in different age groups and found significant results in the adolescent age group (between 18-24). This signified that adolescents were at a higher risk of developing COVID-19 infection as compared to other age groups.

### c.) Implications of all the available evidence

By providing as assessment of prevalence of COVID-19 in vaping adolescent population, our study provides unique information for clinicians and patients about the importance of avoidance from vaping in the adolescent population. As more prospective trials are performed with respect to COVID-19 and vaping, more information will be ascertained to enhance our understanding impacts of both novel diseases.

## Results

Our study included an analysis of a population of 328,300,544 people across the US that was sampled from 2019 estimated population per combined data provided by the US Census Department and Worldometers (7,8) (Table 1). A total of 1,067,508 cases of coronavirus were identified across 50 states and the District of Columbia as per the data investigated Worldometers up until 04/30/2020 (7,8) (Table 1). The prevalence was divided into 4 distinct categories by age group, as noted in the Methods section. The Spearman correlation analysis demonstrated that persons vaping between 18 years and 24 years of age had a correlation coefficient of 0.278 with prevalence of COVID-19 infection (p=0.048). Vaping high school students (age 14-18) had a correlation coefficient of 0.153 with prevalence of COVID-19 (p=0.328). Persons vaping in the age group 25-45 years had a correlation coefficient of 0.101 in association to COVID-19 prevalence (p=0.478). And finally, persons vaping between the age 45-65 years old had a correlation coefficient 0.130 with respect to COVID-19 prevalence (p=0.364) (Table 2).

**Table 1.** Prevalence of COVID-19 cases and Vaping in Late April 2020.

| State | Cases | Estimated<br>2019<br>Population | Cases/100k | E-cigarette use by age group<br>(%) |  |  |  |
| --- | --- | --- | --- | --- | --- | --- | --- |
|  |  |  |  | High<br>School | 18-24<br>years | 25-44<br>years | 45-65<br>years |
| Alabama | 7088 | 4,903,185 | 0.006918 | 24.5 | 8.5 | 6.6 | 4.4 |
| Alaska | 355 | 731,545 | 0.020607 | 15.7 | 18.9 | 7.4 | 2.3 |
| Arizona | 7655 | 7,278,717 | 0.009508 | 16.1 | 12.5 | 6.6 | 4 |
| Arkansas | 3281 | 3,017,825 | 0.009198 | 13.9 | 15.3 | 10.3 | 4.8 |
| California | 50130 | 39,512,223 | 0.007882 | 17.3 | 6.4 | 4.8 | 1.2 |
| Colorado | 15284 | 5,758,736 | 0.003768 | 26.2 | 17.6 | 6.8 | 2.9 |
| Connecticut | 27700 | 3,565,287 | 0.001287 | N/A | 8.7 | 7.8 | 2.7 |
| Delaware | 4734 | 973,764 | 0.002057 | 13.6 | 10.3 | 7.7 | 2.7 |
| District Of<br>Columbia | 4323 | 705,749 | 0.001633 | 10.9 | 5.8 | 2.1 | 2.1 |
| Florida | 33690 | 21,477,737 | 0.006375 | N/A | 19.5 | 8.7 | 3.6 |
| Georgia | 26264 | 10,617,423 | 0.004043 | N/A | 13.6 | 6.9 | 3.1 |
| Hawaii | 618 | 1,415,872 | 0.022911 | 25.5 | 23.8 | 9.4 | 3.7 |
| Idaho | 1984 | 1,787,147 | 0.009008 | 14.3 | 20.8 | 6.8 | 3.4 |
| Illinois | 52918 | 12,671,821 | 0.002395 | 13.2 | 10.3 | 5.6 | 3.1 |
| Indiana | 17835 | 6,732,219 | 0.003775 | 23.9 | 17.5 | 8.3 | 4.6 |
| Iowa | 7145 | 3,155,070 | 0.004416 | 9.0 | 16.6 | 6.1 | 3.1 |
| Kansas | 4413 | 2,913,314 | 0.006602 | 10.6 | 9.8 | 6.1 | 3.2 |
| Kentucky | 4708 | 4,467,673 | 0.00949 | 14.1 | 13.1 | 7 | 5.9 |
| Louisiana | 28001 | 4,648,794 | 0.00166 | 12.2 | 14.8 | 7.8 | 3.1 |
| **Maine** | **1095** | 1,344,212 | 0.012276 | 15.8 | 9.7 | 6.5 | 2.7 |
| **Maryland** | **21742** | 6,045,680 | 0.002781 | 13.3 | 11.3 | 6 | 2.6 |
| **Massachusetts** | **62205** | 6,949,503 | 0.001117 | 20.1 | 13 | 8.2 | 3.6 |
| **Michigan** | **41379** | 9,986,857 | 0.002414 | 14.8 | 12.8 | 6 | 3.4 |
| **Minnesota** | **5136** | 5,639,632 | 0.010981 | N/A | 18.1 | 5.7 | 2.5 |
| **Mississippi** | **6815** | 2,976,149 | 0.004367 | 22.9 | 11.7 | 8.6 | 3.5 |
| **Missouri** | **7818** | 6,137,428 | 0.00785 | 10.9 | 14.4 | 7.6 | 3.9 |
| **Montana** | **453** | 1,068,778 | 0.023593 | 22.5 | 15 | 6.7 | 2.3 |
| **Nebraska** | **4281** | 1,934,408 | 0.004519 | 9.4 | 15.4 | 6.7 | 3.5 |
| **Nevada** | **5053** | 3,080,156 | 0.006096 | 15.5 | 12.4 | 7.1 | 2.9 |
| **New Hampshire** | **2146** | 1,359,711 | 0.006336 | 23.8 | 11.1 | 8 | 3 |
| **New Jersey** | **118652** | 8,882,190 | 0.000749 | N/A | 11.5 | 6.4 | 2.5 |
| **New Mexico** | **3411** | 2,096,829 | 0.006147 | 24.7 | 12 | 7 | 2.5 |
| **New York** | **304372** | 19,453,561 | 0.000639 | 14.5 | 14.3 | 7.3 | 3 |
| **North Carolina** | **10754** | 10,488,084 | 0.009753 | 22.1 | 15.9 | 6.7 | 3.3 |
| **North Dakota** | **1067** | 762,062 | 0.007142 | 20.6 | 23 | 6.6 | 2.6 |
| **Ohio** | **18027** | 11,689,100 | 0.006484 | N/A | 13.8 | 7.3 | 3.8 |
| **Oklahoma** | **3618** | 3,956,971 | 0.010937 | 16.4 | 15.4 | 8.5 | 5.3 |
| **Oregon** | **2510** | 4,217,737 | 0.016804 | N/A | 11.6 | 8.5 | 3.5 |
| **Pennsylvania** | **47971** | 12,801,989 | 0.002669 | 11.3 | 7.8 | 6.8 | 4.3 |
| **Rhode Island** | **8621** | 1,059,361 | 0.001229 | 20.1 | 11.3 | 6.9 | 4.6 |
| **South Carolina** | **6095** | 5,148,714 | 0.008447 | 11.9 | 9.1 | 5.5 | 2.9 |
| **South Dakota** | **2449** | 884,659 | 0.003612 | 17.3 | 19.6 | 4.5 | 2 |
| **Tennessee** | **10735** | 6,833,174 | 0.006365 | 11.5 | 16.1 | 7.7 | 3.5 |
| **Texas** | **28727** | 28,995,881 | 0.010094 | 10.3 | 12.9 | 6.7 | 3.7 |
| Utah | **4672** | 3,205,958 | 0.006862 | 7.6 | 14.6 | 7.1 | 3 |
| Vermont | **866** | 623,989 | 0.007205 | 12.0 | 6.1 | 5.9 | 1.3 |
| Virginia | **15847** | 8,535,519 | 0.005386 | 11.8 | 13.7 | 6.7 | 2.7 |
| Washington | **14327** | 7,614,893 | 0.005315 | N/A | 9.3 | 5.3 | 3.5 |
| West Virginia | **1125** | 1,792,065 | 0.015929 | 14.3 | 10.9 | 9.1 | 4.2 |
| Wisconsin | **6854** | 5,822,434 | 0.008495 | 11.6 | 16.9 | 5.5 | 2.5 |
| Wyoming | **559** | 578,759 | 0.010353 | 29.6 | 21.8 | 7 | 4 |

**Table 2.** Correlations Between Vaping Rate and COVID-19 Prevalence in Late April 2020.

| Age range | Correlation coefficient | P value |
| --- | --- | --- |
| High School | 0.153 | 0.328 |
| 18-24 years | **0.278** | **0.048** |
| 25-45 years | 0.101 | 0.478 |
| 45-65 years | 0.130 | 0.364 |

## Discussion

Over 320 million people across the US were sampled from 2019 estimated population per combined data provided by the US Census Department and Worldometers (7,8). Approximately 1 million cases of COVID-19 were identified across 50 states and the District of Columbia as per the data investigated via Worldometers up until 04/30/2020. The vast number of cases involved in our study appreciably increases the power of our study. More importantly, we observed statistically significant results in the adolescent age group with relation to vaping and COVID-19 prevalence. Our study demonstrated that persons vaping between 18 years and 24 years of age were at a slightly higher risk of developing COVID-19 infection. High school students (age 14-18), 25-45 year old persons, and 45-65 year old persons did not illustrate a statistically significant relationship between vaping and COVID-19 infection as the p-values of the Spearman analysis were greater than 0.05. Our current analysis from the data obtained via Worldometers shows high concentration of vaping prevalence among 18-24 year-old persons in North Dakota, Wyoming, Idaho, and Florida; other high concentration areas for the other age groups may be identified in Figure 1.

**Figure 1.**
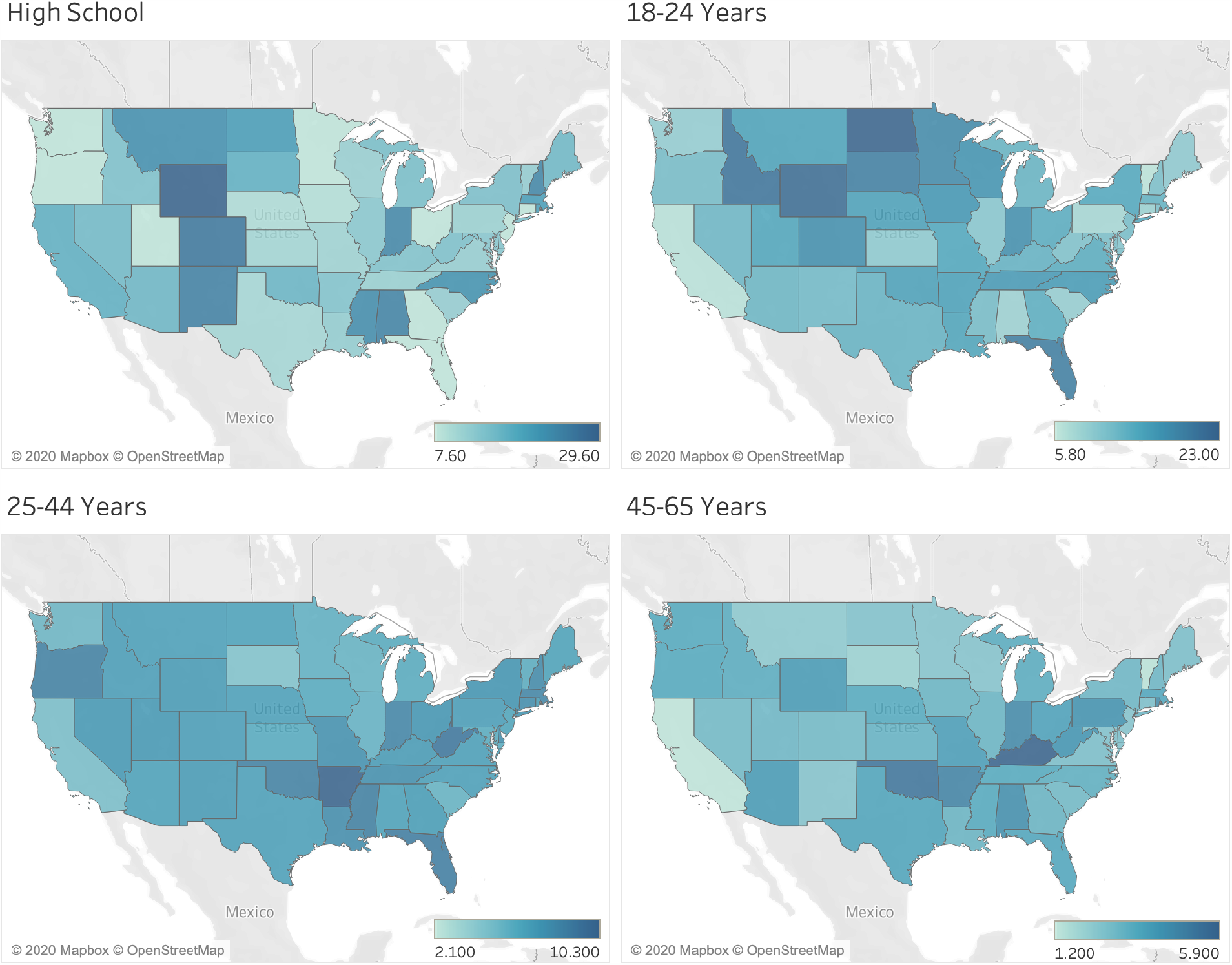
Vaping prevalence map for high school students, 18-24 year old persons, 25-44 year old persons, and 45-65 year old persons.

COVID-19 has been shown to be associated with higher morbidity and mortality in patients with chronic medical conditions. Diabetes mellitus, chronic lung disease (including asthma, chronic obstructive pulmonary disease and emphysema), and cardiovascular disease were noted to be the most common comorbidities associated with severe outcomes (9). Former smoking and current smoking were both lower on the table of risk factors that may be associated with severe outcomes (9). However, at this juncture, there are currently no data or findings suggesting a correlation between vaping and COVID-19 (10). Most of the aforementioned conditions lead to higher propensity of deaths in the elderly population. Death rates have been recorded as high as approximately 3.5%, 8.0%, and 15% in the 6th, 7th and 8th decades of life, respectively (11).

Contrast the elderly death rate with the death rate in the young adult population, where the estimated death rate between 18-year-old persons to 24-year-old persons that are infected with COVID-19 is less than 0.5% (11). While our study illustrates the correlation between vaping in 18-24 year old persons and COVID-19 prevalence may be weakly positive at 0.278, the results are significant enough to raise certain concerns. Complications from past and current cigarette smoking are known and former smokers and current smokers are at risk of severe complications from COVID-19 infection. In a systematic review of COVID-19 and smoking, 5 studies were used to demonstrate former and current smokers were 1.4 times more likely to suffer severe complications from COVID-19 infection as compared to nonsmokers (12). From current studies, vaporizers are known to add an additional component of risk of significant pulmonary complications. A study involving 98 patients with median age 21 years old demonstrated 95% of patients were hospitalized and approximately 26% of patients underwent intubation and mechanical ventilation (13). Our study demonstrates that the addition of vaporizers may put our otherwise healthy adolescent population at higher risk of COVID-19 infection, which may increase the propensity for morbidity and mortality.

Potential limitations were observed while conducting this observational study. Because the data obtained was from public domain, we did not have access to other health information regarding all of the patients that were diagnosed with COVID-19. Potential comorbidities, and thus potential confounders, were unable to be identified as this information was not obtainable via the public domain. This may limit the interpretation of the results. We explain our findings cautiously and hope that further prospective studies may be performed in order to evaluate the strength of the conclusion presented in this manuscript.

## Conclusion

Increased COVID-19 prevalence is associated with vaping in the adolescent population between ages 18 and 24. As we continue to learn more about COVID-19, it is imperative that we abate additional risk factors that may place persons, especially our younger population, at risk of this deadly viral infection. Further prospective studies should be performed in order investigate the severity of outcomes of electronic cigarette use in association with COVID-19 infection.

## Data Availability

All data included in this study is available in the public domain. Thus, there is no limitation in obtaining and reviewing the data provided in this manuscript.

## Acknowledgements

The authors would like to thank the Southern Illinois University School of Medicine Department of Internal Medicine for their support.

## Author Contributions

All authors contributed equally in the preparation of this manuscript.

## Conflicts of Interest

Nothing to disclose.

## Funding Source

Nothing to disclose. We had full access to all of the resources through public domain.

## Notes

### Competing Interest Statement

The authors have declared no competing interest.

### Funding Statement

There are no funding sources to disclose.

### Author Declarations

No direct patient care or contact involved in this observational study thus the study was exempt from IRB oversight. Institution Review Board: SCRIHS.

## References

1. “Cases in U.S.” Centers for Disease Control and Prevention, 6 June 2020. http://www.cdc.gov/coronavirus/2019-ncov/cases-updates/cases-in-us.html#1.

2. “COVID-19 Dasboard by the Center for Systems Science and Engineering (CSSE) at Johns Hopkins University (JHU)”. Coronavirus Resource Center, Johns Hopkins University and Medicine. 6 June 2020. https://coronavirus.jhu.edu/map.html

3. Shereen MA, Khan S, Kazmi A, Bashir N, Siddique R. “COVID-19 infection: Origin, transmission, and characteristics of human coronaviruses”. J Adv Res. 16 Mar 2020;24:91–98. doi: 10.1016/j.jare.2020.03.005. PMID: 32257431; PMCID: PMC7113610.

4. He X, Lau EHY, Wu P, et al. “Temporal Dynamics in Viral Shedding and Transmissibility of COVID-19.” MedRxiv, Cold Spring Harbor Laboratory Press, 1 Jan. 2020, doi.org/10.1101/2020.03.15.20036707.

5. Wang J, Du G. COVID-19 may transmit through aerosol. Ir J Med Sci (2020). https://doi.org/10.1007/s11845-020-02218-2

6. Evans-Polce RJ, Patrick ME, Lanza ST, Miech RA, O’Malley PM, Johnston LD. “Reasons for Vaping Among U.S. 12th Graders”. J Adolesc Health. 2018 Apr;62(4):457–462. doi: 10.1016/j.jadohealth.2017.10.009. Epub 2017 Dec 20. PMID: 29273302; PMCID: PMC5866738.

7. Protano C, Avino P, Manigrasso M, et al. “Environmental Electronic Vape Exposure from Four Different Generations of Electronic Cigarettes: Airborne Particulate Matter Levels”. Int. J. Environ. Res. Public Health 2018, 15, 2172. https://doi.org/10.3390/ijerph15102172

8. US Census Bureau. “State Population Totals: 2010-2019.” The United States Census Bureau, 30 Dec. 2019, www.census.gov/data/tables/time-series/demo/popest/2010s-state-total.html.

9. “State Tobacco Activities Tracking and Evaluation (STATE) System.” Centers for Disease Control and Prevention, Centers for Disease Control and Prevention, 18 Mar. 2020, www.cdc.gov/statesystem/index.html.

10. “Preliminary Estimates of the Prevalence of Selected Underlying Health Conditions Among Patients with Coronavirus Disease 2019 - United States, February 12–March 28, 2020.” Centers for Disease Control and Prevention, Centers for Disease Control and Prevention, 2 Apr. 2020, www.cdc.gov/mmwr/volumes/69/wr/mm6913e2.htm.

11. Ducharme, J. “Is There Actually a Link Between Vaping and COVID-19?” Time, 23 Mar. 2020, time.com/5807214/vaping-coronavirus/.

12. “Age, Sex, Existing Conditions of COVID-19 Cases and Deaths.” Worldometer, www.worldometers.info/coronavirus/coronavirus-age-sex-demographics/.

13. Vardavas CI, Nikitara K. “COVID-19 and smoking: A systematic review of the evidence.” Tobacco induced diseases vol. 18 20. 20 Mar. 2020, doi:10.18332/tid/119324

14. Layden JE, Ghinai I, Pray I, et al. Pulmonary Illness Related to E-Cigarette Use in Illinois and Wisconsin - Final Report. N Engl J Med 2020;382(10):903–916. doi:10.1056/NEJMoa1911614

